# VIDAS^®^ TB-IGRA accuracy in tuberculosis patients and persons at varying risk of exposure

**DOI:** 10.1101/2024.07.03.24309158

**Authors:** Delia Goletti, Niaz Banaei, Rahul Batra, Anne Emmanuelle Berger, Azra Blazevic, Elisabeth Botelho-Nevers, Ronan Breen, Natalie Bruiners, Emmanuelle Cambau, Etienne Carbonnelle, Charles L. Daley, Cécile Descotes-Genon, Francesco Di Gennaro, Florence Doucet-Populaire, Aliasgar Esmail, Julia Dolores Estrada Guzman, Luc Fontana, Maria Laura Gennaro, Deborah Handler, Rosa María Herrera Torres, Daniel Hoft, Nahed Ismail, Margaux Isnard, Alfred Lardizabal, François Xavier Lesage, Amanda Lopes, Williams Luciano López Vidal, Rene Machado Contreras, Philippe Manivet, Hubert Marotte, Frédéric Méchaï, Amel Medjahed-Artebasse, Richard Meldau, Yves Mérieux, Jacques Morel, Faiza Mougari, Suzette Oelofse, Fabrizio Palmieri, Jean Luc Perrot, Elisa Petruccioli, David T. Pride, Edouard Tuaillon, Caryn Upton, Naadira Vanker, Keertan Dheda

## Abstract

**Background:** Detection and treatment of individuals with presumed latent tuberculosis (TB) infection (i.e., excluding active disease; LTBI) is imperative to achieve global TB control, as they represent a potential transmission reservoir. However, more sensitive and user-friendly diagnostic tools are needed.

**Methods:** We evaluated the accuracy for TB infection detection of the new VIDAS^®^ TB-IGRA (bioMérieux), a fully automated, single tube (thus eliminating the need for batch testing) overnight incubation assay, compared to the QuantiFERON^®^-TB Gold Plus (QFT-Plus, QIAGEN), in a global multi-centre cross-sectional study (NCT04048018) that included patients with TB disease (*n*=200) or participants at varying levels of TB exposure (*n*=1460; mixed exposure-risk-population).

**Results:** VIDAS^®^ TB-IGRA identified TB disease with greater sensitivity than QFT-Plus (97.5% *vs*. 80.7%, *P*<0.01%), and yielding significantly fewer false-negatives (2.5% *vs*. 17.5%; *P*<0.01%) and indeterminate results (1.0% *vs*. 9.5%; *P*=0.02%). In the mixed exposure-risk-population, negative (NPA) and positive percent agreement (PPA) were 90.1% (1097/1217) and 92.1% (223/242), respectively. PPA increased with TB-exposure risk (up to 95.7% for high-risk participants), whereas NPA decreased (starting from 96.9% for low-risk participants). Regression analyses revealed that VIDAS^®^ TB-IGRA had a better fit with the risk-exposure gradient than the QFT-Plus. Specificity in extremely low TB-exposure risk participants (*n* = 125) was high for both VIDAS^®^ TB-IGRA and QFT-Plus (97.6% *vs*. 95.2%; *P*=8.33%).

**Conclusions:** VIDAS^®^ TB-IGRA displayed greater sensitivity than QFT-Plus, had a lower indeterminate rate, correlated better with an exposure gradient, and was highly specific, suggesting that it is a potentially valuable tool for the diagnosis of LTBI.

**Take-home message:** The first fully automated interferon-γ-release assay—the bioMérieux VIDAS^®^ TB-IGRA—is highly specific and displays greater sensitivity than QuantiFERON^®^-TB Gold Plus, and thus represents a valuable new and streamlined diagnostic tool for TB infection.

## INTRODUCTION

Tuberculosis (TB), due to infection by *Mycobacterium tuberculosis* (*Mtb*) is a significant global health problem that resulted in 1.3 million estimated deaths in 2022 [1]. TB is transmitted from person-to-person *via* bacteria-containing respiratory droplets [2]. The probability of being infected depends on the likelihood and degree of exposure, which is referred to as TB-exposure risk [3, 4]. Approximately 5–10% of *Mtb*-infected individuals develop TB disease (also called active TB); however, most infections (90–95%) are controlled by the host immune response and do not progress [2]. This asymptomatic non-transmissible state, known as latent TB infection (LTBI), can later progress to TB disease in response to triggers, such as impaired immune function, and represents an important reservoir for disease. Current research further suggests a continuum from exposure to disease, with transition between states dependent on modulators of host immunity [2, 5–9]. Hereafter the term TB infection is used to represent all stages of infection with *M. tuberculosis* without clinical manifestations of TB disease, as suggested in [10].

TB diagnosis remains challenging, particularly in middle/low-income countries. Furthermore, there is no gold-standard for the diagnosis of TB infection. Current methods –the tuberculin skin test (TST) and interferon (IFN)-γ-release assays (IGRAs)– assess host response to infection [9, 11–14] and neither can differentiate between TB disease and TB infection or predict risk of progression to TB disease. IGRAs, such as QuantiFERON-TB Gold Plus (QFT-Plus; QIAGEN) and T-SPOT-TB (Oxford Immunotec), are *in vitro* blood tests that measure T cell-mediated release of IFN-γ in response to synthetic *Mtb* complex-specific antigens (*e.g.,* ESAT-6 and CFP-10) [15]. These tests show enhanced specificity for *Mtb* relative to the TST and can be performed in a single patient visit.

Studies have shown the benefit of systematic testing and treatment for presumed TB infection in people with HIV, pulmonary-TB household contacts under-5 years old and groups at clinical risk for TB, including immigrants to low-burden settings, those receiving anti-tumour necrosis factor (TNF) treatment or dialysis, and organ-related or bone marrow transplant patients [16–18]. More recently, the World Health Organization (WHO) has advocated for TB infection screening and treatment of household contacts, even in TB-endemic settings, and modelling studies have indicated TB eradication will be impossible without treating the large reservoir of TB infection [19]. These findings highlight the importance of streamlined testing methods for TB infection. However, currently available IGRAs are often cumbersome to perform, requiring several steps and often the need for batch testing, which leads to complex laboratory workflows, introduces variability, and delays results [11, 12, 15, 20].

To address the unmet need for improved TB testing methods, bioMérieux developed the VIDAS^®^ TB-IGRA, the first IGRA to be fully automated, from sample pipetting to result interpretation. This test, based on ESAT-6 and CFP-10 peptides-specific CD4^+^ and CD8^+^ T-cell responses [21] requires a single whole-blood sample per patient and provides results within 17 hours. Here, we assessed the accuracy for TB infection detection of VIDAS^®^ TB-IGRA *vs*. QFT-Plus in an international, multi-centre, cross-sectional study of participants with culture-confirmed TB disease or with varying levels of TB-exposure risk (mixed exposure risk population). Since there is no gold standard to diagnose LTBI, sensitivity was evaluated in patients with TB disease and specificity was evaluated in persons at extremely low TB risk. Figure 1 provides an overview of the study sub-groups and the performance characteristics.

**Figure 1.**
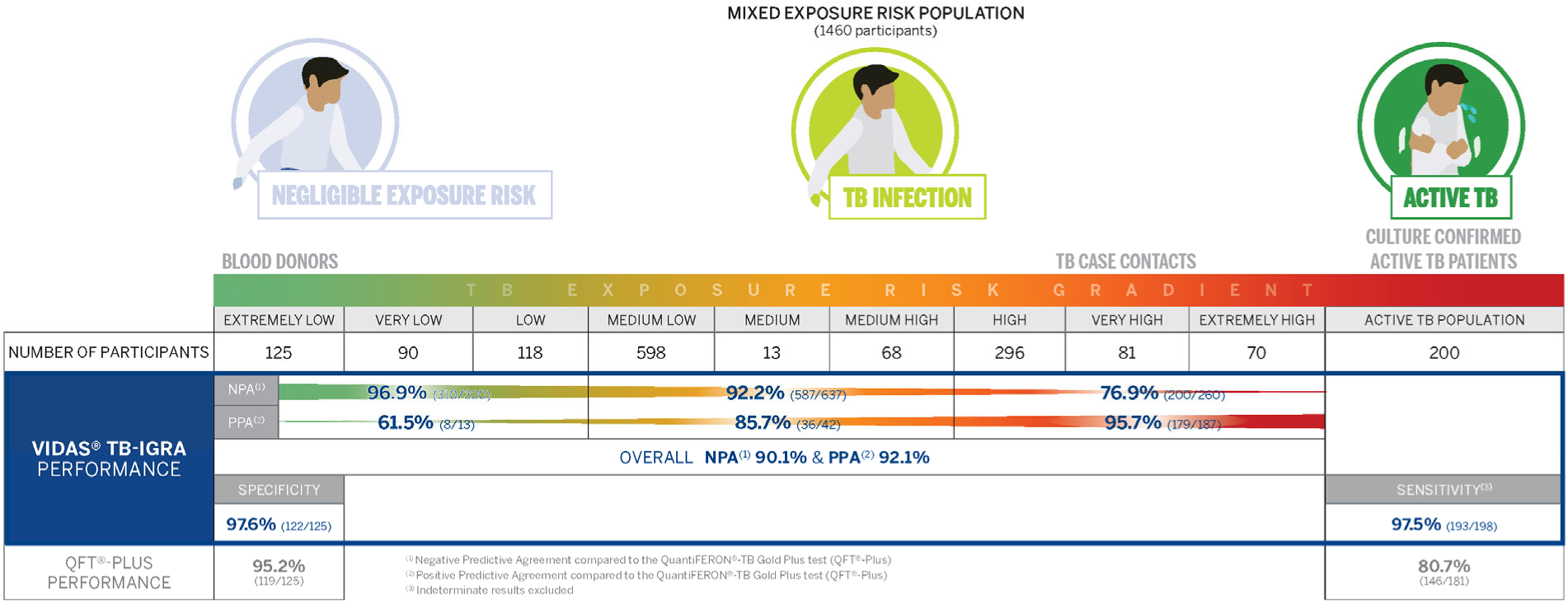
Overview of the study populations and the accuracy evaluated for VIDAS^®^ TB-IGRA and QFT-Plus.

## METHODS

### Study Design and Participants

Study participants were recruited, between December 2019 and December 2020, incorporating i) the TB disease population, or ii) the mixed exposure risk population. The TB disease population contained 200 culture-confirmed patients with TB disease recruited at 10 sites in both low (France, Italy, UK, USA) and high TB-incidence countries (South Africa, Mexico). The mixed exposure risk population contained 1,460 individuals recruited from 18 sites, with or without identified risk factors for TB, and no history of TB disease or TB infection, as assessed by questionnaire. See Supplementary Table S1 and Supplementary Figure S2 for more details on study sites and participant flow, respectively.

The mixed exposure risk population was stratified in nine subgroups, along a gradient of TB-exposure risk, ranging from extremely low (blood-bank donors in France) to extremely high risk (same-bedroom TB-case contacts). The subgroups were further pooled into aggregate low-, medium-, and high-risk groups (Supplementary Table S2).

In this paper, the term TB infection is used to represent all stages of infection with *Mtb* without clinical manifestations of TB disease, as suggested in [10].

Patients with TB disease were recruited within 15 days of TB treatment. TB disease was defined as positive TB culture followed by *Mtb*-complex species identification. HIV-infected patients were excluded. In addition, the following exclusion criteria applied to both populations: treatment with TNF-α inhibitors within 3 months prior to enrolment, TST within 12 weeks prior to enrolment, self-reported pregnancy, history of nontuberculous mycobacterial infection. Patient demographic information is listed in table 1.

**TABLE 1.** Demographic information for the participants enrolled in this study.

|  |  | **TB disease**<br>**population**<br>**(*n* = 200)** | **Mixed exposure risk**<br>**population (*n* = 1,460)** | **P-value \*** |
| --- | --- | --- | --- | --- |
| Gender<br>Number (%) | Female | 57 (28.5%) | 947 (64.9%) | < 0.01% |
|  | Male | 143 (71.5%) | 513 (35.1%) |  |
| Age | IQR | 19 | 23 | 0.03% |
|  | Median | 31 | 35 |  |
| BCG-vaccinated<br>Number (%) | Yes | 132 (66.0%) | 534 (36.6%) | < 0.01% |
|  | No | 16 (8.0%) | 627 (42.9%) |  |
|  | Unknown | 52 (26.0%) | 299 (20.5%) |  |
| HIV-infected***<br>Number (%) | Yes | N/A*** | 22 (1.5%) | N/A |
|  | No | N/A*** | 1210 (82.9%) |  |
|  | Unknown | N/A*** | 228 (15.6%) |  |

\* TB disease and Mixed exposure risk populations were compared either with the Mann-Whitney test for Age, or with the Chi square test for Gender and BCG vaccination.
\*\* HIV-positivity was an exclusion criterion for patients in the TB disease population.
**Abbreviations:** BCG, bacille Calmette–Guérin; HIV, human immunodeficiency virus; IQR, interquartile range; N/A, Not Available.

Written informed consent was obtained from all participants. Study protocols were approved by Institutional Review Boards (IRB) or Independent Ethics Committees (IEC) as appropriate (Supplementary Table S3). See supplementary information for the full list of study sites and site staff.

### Sample collection and testing

Whole blood samples from each study participant were tested with the VIDAS^®^ TB-IGRA (bioMérieux, Marcy-l’Étoile, France) and QFT-Plus (QIAGEN, Hilden, Germany). Testing procedures and result analysis followed the manufacturer’s instructions.

The VIDAS^®^ TB-IGRA was performed with reagents and VIDAS^®^3 instruments provided by bioMérieux. For this test, samples are loaded into the instrument with three stimulation reagents: AG, antigen; MIT, mitogen (positive control); NIL, negative control. All steps are performed by the VIDAS^®^3 instrument, i.e., sample and reagent transfers, 37°C incubation for 16 hours, automated enzyme-linked immunofluorescent assay (ELFA) to measure IFN-γ. Qualitative results are automatically determined using previously established thresholds (Supplementary Table S4), notably 0.35 IU/mL for AG-NIL values.

### Statistical analyses

The rate of positive, negative or indeterminate results was calculated relative to the total number of results for the population analysed, i.e., including indeterminate results. Statistical significance of differences between QFT-Plus and VIDAS^®^ TB-IGRA was calculated using McNemar’s test for paired data, with *P*<5% considered as significant.

For the TB disease population, sensitivity of QFT-Plus and VIDAS^®^ TB-IGRA were independently calculated, excluding indeterminate results.

For the mixed exposure risk population, positive percent agreement (PPA) and negative percent agreement (NPA) of VIDAS^®^ TB-IGRA relative to QFT-Plus were calculated, along with 95% confidence intervals (95% CIs). Study participants with indeterminate results from either test were not included in NPA/PPA computations. See Supplementary Methods for more information.

Logistic regression was performed to assess association between TB-exposure levels and positivity rates for individuals in the mixed exposure risk population (Table 5) considering: 1) the three aggregate risk groups (low, medium, and high) and 2) the three subgroups within the aggregate high-risk group (i.e., high-, very high-, and extremely high-risk subgroups). In each case, a univariate regression (unadjusted model) was performed. When necessary, the model was adjusted using three potential confounding variables that were present for all individuals and potentially related to TB infection: age, sex, and geographic area (defined as EU, US, or rest of world). For each variable, independence from exposure-risk level was first assessed using the Chi-square test for qualitative variables (sex and geographic area) and the Kruskal–Wallis Test for continuous variables (age), with α=1% for independence tests, before including it as a confounding factor in the model. This avoids sampling over-representation of some items in the different exposure-risk levels. Results are reported as unadjusted and adjusted odds ratios (OR) with 95% CIs. All statistical analyses were performed using SAS software v.9.4.

## RESULTS

### VIDAS^®^ TB-IGRA showed superior sensitivity in the TB disease population

In the TB disease population (200 culture-confirmed TB disease patients) the VIDAS^®^ TB-IGRA showed higher sensitivity compared to QFT-Plus (97.5% *vs.* 80.7% respectively, *P*<0.01%, see table 2). VIDAS^®^ TB-IGRA significantly detected more TB disease cases than QFT-Plus (193 vs 146 among the 200 patients included in the analysis, i.e., 96.5% *vs*. 73.0%, *P*<0.01%), and led to fewer false-negative (2.5% *vs.* 17.5%, P<0.01%) and indeterminate results (1.0% *vs.* 9.5%, P=0.02%). In summary, patients with TB disease were better identified by VIDAS^®^ TB-IGRA as compared to QFT-Plus.

**TABLE 2.** Results obtained with the VIDAS^®^ TB-IGRA and QFT-Plus tests for the TB disease and mixed exposure risk populations, as well as the blood donor subgroup at extremely low risk for TB. Percentages are shown in brackets while 95% Confidence Intervals are shown in square brackets.

|  | TB disease population (*n* = 200) |  |  |
| --- | --- | --- | --- |
|  | VIDAS® TB-IGRA | QFT-Plus | *P*-value** |
| **True Positive** | 193 (96.5%) | 146 (73.0%) | <0.01% |
| **False Negative** | 5 (2.5%) | 35 (17.5%) | <0.01% |
| **Indeterminate** | 2 (1.0%) | 19 (9.5%) | 0.02% |
| **Sensitivity\***<br>**[95%CI]** | 97.5% (193/198)<br>[94.2, 99.2]% | 80.7% (146/181)<br>[74.3, 85.8]% | <0.01%*** |
|  | Mixed exposure risk population (*n* = 1,460) |  |  |
|  | VIDAS® TB-IGRA | QFT-Plus | *P*-value** |
| **Positive** | 343 (23.5%) | 242 (16.6%) | <0.01% |
| **Negative** | 1,117 (76.5%) | 1,217 (83.4%) | <0.01% |
| **Indeterminate** | 0 (0.0%) | 1 (0.1%) | NC |
|  | Extremely Low Risk Subgroup / Blood Donors (*n* = 125) |  |  |
|  | VIDAS® TB-IGRA | QFT-Plus | *P*-value |
| **Positive** | 3 (2.4%) | 6 (4.8%) | NC |
| **Negative** | 122 (97.6%) | 119 (95.2%) | 8.33% (NS) |
| **Indeterminate** | 0 (0.0%) | 0 (0.0%) | NA |
| **Specificity**<br>**[95%CI]** | 97.6% (122/125)<br>[93.1, 99.5]% | 95.2% (119/125)<br>[89.8, 98.2]% | 8.33% (NS) |

\* Excluding indeterminate results.
\*\* Unless otherwise stated, the McNemar's test for paired data was used, comparing the results identified on the row with all other results, e.g., for the first row: True Positive vs (False Negative + Indeterminate).
\*\*\* Calculated using the Bowker's Symmetry Test.
**Abbreviations:** CI, confidence interval. NA, not applicable. NC, not calculated. NS, non-significant. QFT-Plus, QuantiFERON-TB Gold Plus.

### Clinical performance of the VIDAS^®^ TB-IGRA in the mixed exposure risk population

In the mixed exposure risk population (1,460 individuals without TB symptoms, at various levels of TB-exposure risk) the positivity rate was significantly higher for VIDAS^®^ TB-IGRA than QFT-Plus (23.5% *vs*. 16.6% respectively, *P*<0.01%, see table 2). NPA and PPA were 90.1% and 92.1%, respectively (table 3) indicating good agreement between the two tests.

**TABLE 3.** Negative percent agreement (NPA) and positive percent agreement (PPA) for the VIDAS^®^ TB-IGRA relative to the QFT-Plus test for participants in the mixed exposure risk population.

| Mixed exposure risk population |  | QFT-Plus |  |  |
| --- | --- | --- | --- | --- |
|  |  | Negative | Positive | Total* |
| VIDAS® TB-IGRA | Negative | 1097 | 19 | 1116 |
|  | Positive | 120 | 223 | 343 |
|  | Total* | 1217 | 242 | 1459 |
| NPA * | 90.1% |  |  |  |
| [95%CI] | [88.3; 91.7]% |  |  |  |
| PPA * | 92.1% |  |  |  |
| [95%CI] | [88.1; 94.9]% |  |  |  |

\*Excluding indeterminate results.
**Abbreviations:** CI, confidence interval; QFT-Plus, QuantiFERON-TB Gold Plus.

### Analysis of discrepant results in the mixed exposure risk population

To better understand the agreement between VIDAS^®^ TB-IGRA and QFT-Plus in the mixed exposure risk population, we conducted two additional analyses.

We first evaluated whether discordant results were distributed around the cut-off of each test. We found they were distributed along the entire measuring interval, particularly when the VIDAS^®^ TB-IGRA scored positive (Supplementary Figure S2). This indicates that discrepancies are not merely due to variability around the cut-off, and thus may result from other factors, such as different sensitivity of the tests.

Next, we divided the mixed exposure risk population into nine subgroups ranked along a gradient from extremely-low to extremely-high TB-exposure risk. The subgroups may also be distributed into aggregate low-, medium-, and high-risk groups (Supplementary Table S2). Table 4 contains the VIDAS^®^ TB-IGRA and QFT-Plus results for the subgroups and the aggregate groups. For both tests, the positivity rate increased with exposure risk, which is consistent with the expected likelihood for TB infection. However, the positivity rate was significantly higher for VIDAS^®^ TB-IGRA *vs*. QFT-Plus in the aggregate medium- and high-risk groups, but not in the aggregate low-risk group (figure 2). NPA and PPA (VIDAS^®^ TB-IGRA relative to QFT-Plus) varied along the risk gradient. As shown in figure 3 and Supplementary Table S5, PPA increased with the risk, whereas NPA decreased. For the aggregate low-, medium, high-risk groups, NPA was 96.9%, 92.2%, and 76.9%, respectively, while PPA was 61.5%, 85.7%, and 95.7%, respectively.

**Figure 2.**
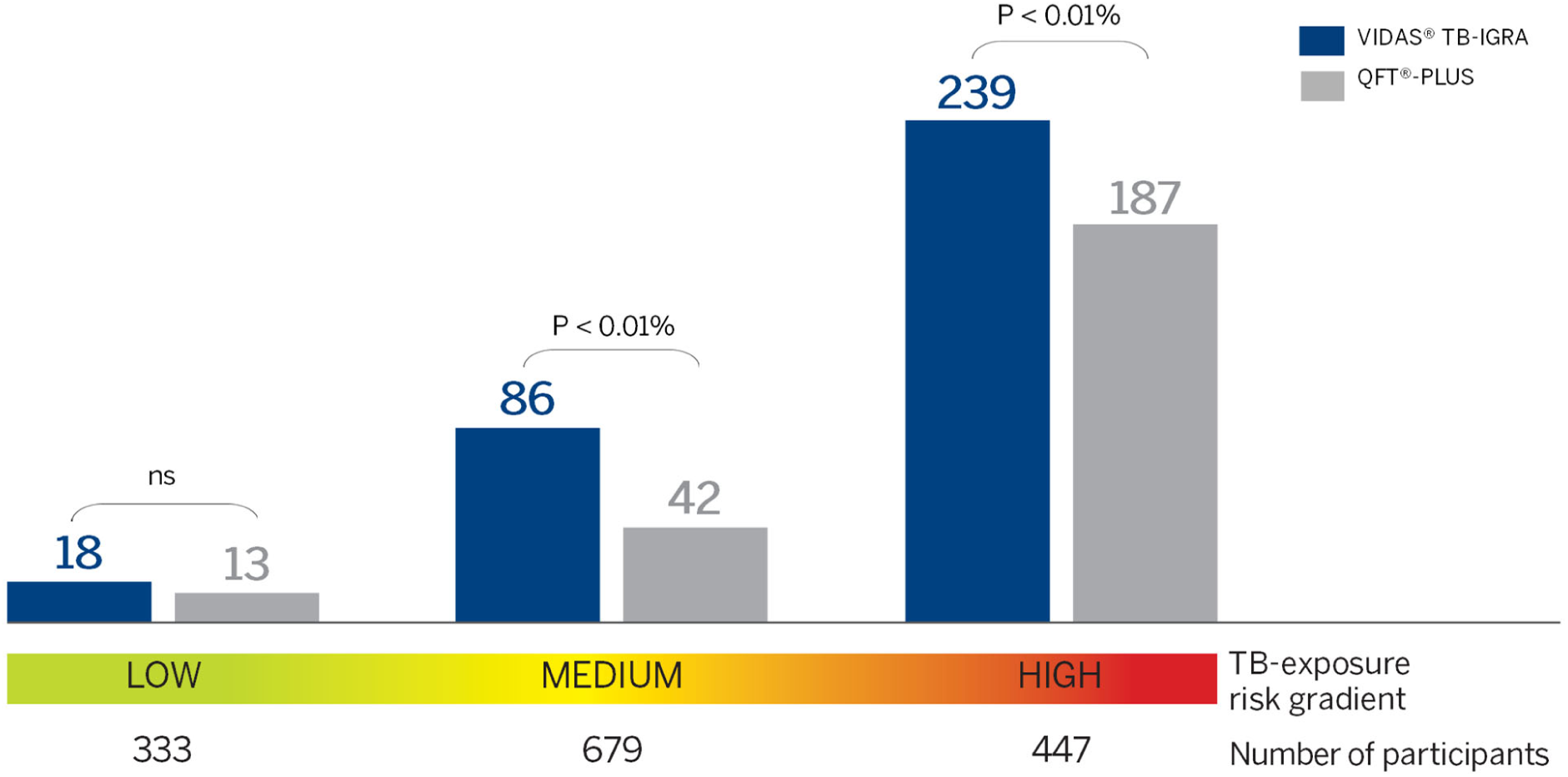
Positive results with VIDAS^®^ TB-IGRA and QFT-Plus for participants in the aggregate low-, medium-, and high-risk groups in the mixed exposure risk population (excludes those with TB disease). ns: not significant; P-values calculated with the McNemar test are shown.

**Figure 3.**
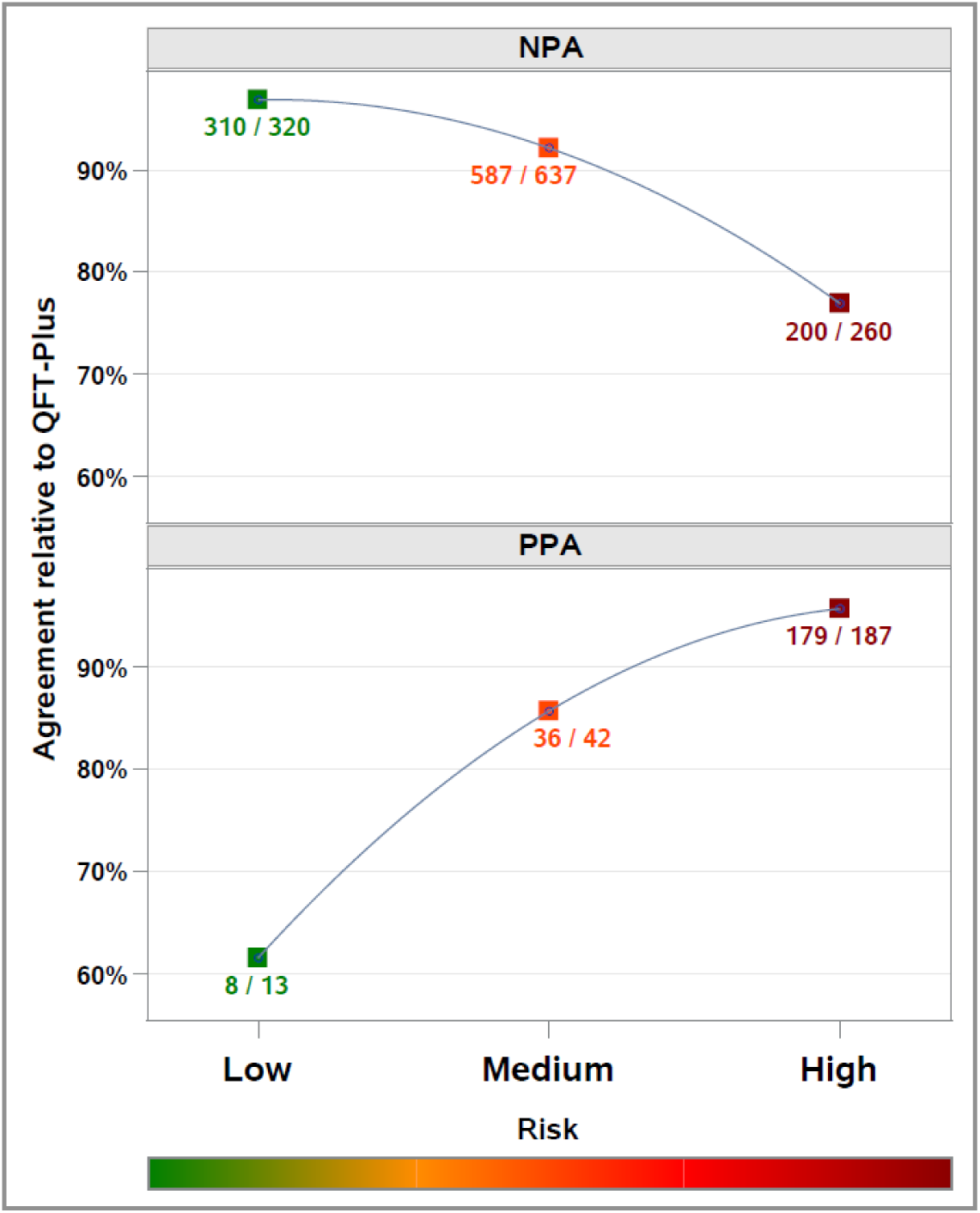
Negative percent agreement (NPA, top graph) and positive percent agreement (PPA, bottom graph) for the VIDAS^®^ TB-IGRA relative to the QFT-Plus for participants in the aggregate low-, medium-, and high-risk groups in the mixed exposure risk population (excludes those with TB disease). Numbers reported next to the points correspond to the number of concordant samples divided by the number of participants in the aggregate group with negative (for NPA) or positive (for PPA) QFT-Plus result. As TB-exposure risk rises, NPA decreases and PPA increases. This indicates that at a low probability of infection, most participants negative by QFT-Plus were also negative by VIDAS^®^ TB-IGRA (high NPA), whereas one-third of those positive by QFT-Plus were negative by VIDAS^®^ TB-IGRA (lower PPA). Conversely, at high probability of infection, most participants positive by QFT-Plus were also positive by VIDAS^®^ TB-IGRA (high PPA), whereas about one-fourth of those negative by QFT were positive by VIDAS^®^ TB-IGRA (lower NPA). PPA and NPA values for all nine TB-exposure risk subcategories are shown in the Supplementary Table S5.

**TABLE 4.** VIDAS^®^ TB-IGRA and QFT-Plus test results for individuals within each subgroup of the mixed exposure risk population.

| TB-Exposure Risk | Participants (n) | Number of Negative Results (%) |  | Number of Positive Results (%) |  |
| --- | --- | --- | --- | --- | --- |
|  |  | VIDAS® TB-IGRA | QFT-Plus | VIDAS® TB-IGRA | QFT-Plus |
| Extremely Low | 125 | 122 (97.6%) | 119 (95.2%) | 3 (2.4%) | 6 (4.8%) |
| Very Low | 90 | 80 (88.9%) | 84 (93.3%) | 10 (11.1%) | 6 (6.7%) |
| Low | 118 | 113 (95.8%) | 117 (99.2%) | 5 (4.2%) | 1 (0.8%) |
| **Aggregate Low** | **333** | **315 (94.6%)** | **320 (96.1%)** | **18 (5.4%)** | **13 (3.9%)** |
| Medium Low | 598 | 553 (92.5%) | 593 (99.2%) | 45 (7.5%) | 5 (0.8%) |
| Medium | 13 | 12 (92.3%) | 13 (100.0%) | 1 (7.7%) | 0 (0.0%) |
| Medium High | 68 | 28 (41.2%) | 31 (45.6%) | 40 (58.8%) | 37 (54.4%) |
| **Aggregate Medium** | **679** | **593 (87.3%)** | **637 (93.8%)** | **86 (12.7%)** | **42 (6.2%)** |
| High | 296 | 152 (51.4%) | 184 (62.2%) | 144 (48.6%) | 112 (37.8%) |
| Very High | 81 | 36 (44.4%) | 45 (55.6%) | 45 (55.6%) | 36 (44.4%) |
| Extremely High | 70 | 20 (28.6%) | 31 (44.3%) | 50 (71.4%) | 39 (55.7%) |
| **Aggregate High** | **447** | **208 (46.5%)** | **260 (58.2%)** | **239 (53.5%)** | **187 (41.8%)** |

**Abbreviations:** QFT-Plus, QuantiFERON-TB Gold Plus.

Moreover, to assess association between exposure-risk level and test positivity, we performed the logistic regression analyses described in the Methods. Among possible confounding variables assessed for individuals within the three aggregate risk categories, only age showed independence from risk-exposure level (*P*=4.4% for age; *P*<0.01% for sex and area) and was therefore analysed by univariate logistic regression. For both tests, univariate odds of a positive result were significantly associated with age (OR:1.01 [95%CI: 1.00, 1.02] for both tests; *P*=1.0% and 2.2%, respectively), indicating that probability of a positive result increased by 1% for each additional year of age. Age was therefore included as a covariate in both adjusted models.

Adjusted and unadjusted ORs for individuals in different TB-exposure risk categories are shown in table 5 and Supplementary Figure S3. For the VIDAS^®^ TB-IGRA, positivity rates of individuals in aggregate medium- and high-risk categories were significantly higher than those in the aggregate low-risk category, with unadjusted ORs of 2.54 [95%CI: 1.5, 4.29] and 20.11 [95%CI: 12.07, 33.49], respectively. This indicates that odds of a positive result with VIDAS^®^ TB-IGRA were 20.11 times higher for individuals in the high-risk *vs*. the low-risk category. For QFT-Plus, only the positivity rate for individuals in the aggregate high-risk category was significantly different from the aggregate low-risk category (unadjusted OR:17.7 [95%CI: 9.86, 31.79]).

**TABLE 5.** Univariate and multivariate odds ratios determined by logistic regression for participants at different levels of TB exposure risk.

|  |  | VIDAS® TB-IGRA |  |  | QFT-Plus |  |  |
| --- | --- | --- | --- | --- | --- | --- | --- |
| Risk group | *n* | Positive Results (%) | Unadjusted OR [95%CI]* | Adjusted OR [95% CI]** | Positive Results (%) | Unadjusted OR [95% CI] | Adjusted OR [95% CI]** |
| Aggregate Low | 333 | 18 (5.4%) | 1 | 1 | 13 (3.9%) | 1 | 1 |
| Aggregate Medium | 679 | 86 (12.7%) | **2.54 [1.50, 4.29]** | **2.55 [1.51, 4.32]** | 42 (6.2%) | 1.62 [0.86, 3.07] | 1.63 [0.86, 3.08] |
| Aggregate High | 447 | 239 (53.5%) | **20.11 [12.07, 33.49]** | **20.17 [12.10, 33.61]** | 187 (41.8%) | **17.70 [9.86, 31.79]** | **17.70 [9.85, 31.81]** |
| Risk subgroup | *n* | Positive Results (%) | Unadjusted OR [95%CI] | Adjusted OR [95% CI]*** | Positive Results (%) | Unadjusted OR [95% CI] | Adjusted OR [95% CI]**** |
| High | 296 | 144 (48.6%) | 1 | 1 | 112 (37.8%) | 1 | 1 |
| Very High | 81 | 45 (55.6%) | 1.32 [0.81, 2.16] | 1.50 [0.86, 2.59] | 36 (44.4%) | 1.31 [0.80, 2.16] | 1.45 [0.84, 2.49] |
| Extremely High | 70 | 50 (71.4%) | **2.64 [1.50, 4.65]** | **2.10 [1.13, 3.91]** | 39 (55.7%) | **2.07 [1.22, 3.50]** | 1.72 [0.97, 3.03] |

\*Significant associations are shown in bold.
\*\*Adjusted for age of study participants.
\*\*\*Adjusted for age of study participants and geographic area (defined as EU, US, or rest of world).
\*\*\*\*Adjusted for geographic area.
**Abbreviations:** CI, confidence interval; OR, odds ratio; QFT-Plus, QuantiFERON Plus.

Results for both tests were similar after adjusting for age; adjusted ORs for aggregate medium- and high-risk groups with VIDAS^®^ TB-IGRA were 2.55 [95%CI: 1.51, 4.32] and 20.17 [95%CI: 12.1, 33.61], respectively, and adjusted OR for aggregate high-risk group with QFT-Plus was 17.7 [95%CI: 9.85, 31.81]. Collectively, these results show a better association between TB-exposure risk and positivity rate with VIDAS^®^ TB-IGRA than QFT-Plus.

We next focused on the subgroups at high-, very high-, and extremely high-risk. For this analysis, all potential cofounding variables (sex, geographic area, and age) showed independence from exposure level (*P*=69.5% for sex; *P*=7.0% for area; *P*=32.5% for age). Univariate analyses revealed significant association between geographic area and positivity rate for both tests (*P*<0.01% for each test); sex was unrelated to positivity rate for both (*P*=12.3% for VIDAS^®^ TB-IGRA; *P*=86.2% for QFT-Plus). Univariate odds of a positive result were also significantly associated with age for the VIDAS^®^ TB-IGRA (OR:1.02 [95%CI: 1.00, 1.03], *P*=1.0 %) but not the QFT-Plus (OR:1.01 [95%CI: 1.00, 1.02], *P*=16.5%). Consequently, age and geographic area were used as cofactors in the VIDAS^®^ TB-IGRA adjusted model, whereas only area was included in the QFT-Plus adjusted model.

Adjusted and unadjusted ORs for the three highest subgroups are shown in table 5 and Supplementary Figure S4. Positivity rates for individuals in the extremely high-risk category were significantly different from those in the high-risk category with both the VIDAS^®^ TB-IGRA and QFT-Plus, with unadjusted ORs of 2.64 [95%CI: 1.5, 4.65] and 2.07 [95%CI: 1.22, 3.5], respectively. However, significance for the QFT-Plus was lost after adjusting for geographic area (OR:1.72 [95%CI: 0.97, 3.03]), whereas for the VIDAS^®^ TB-IGRA, the difference remained significant (OR:2.1 [95%CI: 1.13, 3.91]). These data suggest a better association between exposure and positivity rate for VIDAS^®^ TB-IGRA among TB-case contacts.

### Specificity in blood donors

We evaluated the specificity for TB infection detection by analysing the results obtained from blood donors recruited in Lyon, France (*n*=125), a low endemic area, with 5.7 new cases per 100,000 habitants in 2018 [22]. Because lifestyle and travel history of blood donors are thoroughly analysed (to avoid any health risks for transfusion recipients) they are persons with the lowest TB-exposure risk level. For these individuals, specificity of the VIDAS^®^ TB-IGRA and QFT-Plus was 97.6% and 95.2% (P=8.33%) respectively (table 2), with an NPA of 100% (95% CI: 96.9, 100.0) (Supplementary Table S5). These results indicate that VIDAS^®^ TB-IGRA correctly identifies those not infected with *Mtb* and is therefore a high-specificity test for TB infection.

### Rate of indeterminate results

Lastly, we compared the rate of indeterminate results for the entire study population (*n*=1660) obtained with each test and found that the VIDAS^®^ TB-IGRA yielded significantly fewer indeterminate results than the QFT-Plus: 0.1% (2/1660) *vs*. 1.2% (20/1660), respectively, *P*=0.01%. This suggests the VIDAS^®^ TB-IGRA may be valuable for patient management, as a lower frequency of indeterminate results will decrease the need for repeated testing.

## DISCUSSION

In this study, we assessed the accuracy for TB infection detection of the newly developed VIDAS^®^ TB-IGRA—the first fully automated IGRA—relative to the FDA-approved QFT-Plus, in an international cross-sectional study of culture-confirmed TB patients (TB disease population) or individuals at varying levels of TB-exposure risk (mixed exposure risk population).

In the TB disease population, VIDAS^®^ TB-IGRA showed greater sensitivity than QFT-Plus (97.5% *vs*. 80.7%, respectively) with fewer false-negative and indeterminate results. Notably, the QFT-Plus results are generally consistent with previous studies (11–27). The higher sensitivity of the VIDAS^®^ TB-IGRA may be due to both the reagent design [21] and the full automation of the test.

In the mixed exposure risk population, VIDAS^®^ TB-IGRA yielded a significantly higher positivity rate than QFT-Plus, with NPA and PPA values indicating good agreement between the tests. Then, to assess the accuracy, we stratified this cohort into subgroups with different levels of TB-exposure risk and analysed positivity along the exposure gradient. This is a useful strategy [10] in the absence of a test-based gold standard, as the infection probability depends on likelihood of *Mtb* exposure. For the subgroup at the lowest risk level—individuals expected to be negative—both VIDAS^®^ TB-IGRA and QFT-Plus displayed high specificity. Conversely, for the subgroup at the highest TB-risk level—most likely to be infected—positivity rates for VIDAS^®^ TB-IGRA and QFT-Plus were 71.4% and 55.7%, respectively. These results are consistent with the enhanced sensitivity of the VIDAS^®^ TB-IGRA observed in the TB disease population. NPA and PPA values led to similar conclusions. At the extremes of the TB exposure gradient, both tests largely agree on the most likely result but disagree on the least likely result.

Logistic regression analysis shows that the odds of a positive result with VIDAS^®^ TB-IGRA significantly increased with the risk of TB-exposure for individuals in aggregate medium- and high-risk groups relative to the aggregate low-risk group, even after adjusting for age. For QFT-Plus, only ORs in the aggregate high-risk group were significant (relative to the aggregate low-risk group) before and after adjustment. When focusing only on contact cases (aggregate-high group) we found that odds of a positive result for individuals at the extremely high-risk category were significantly higher than those in the high-risk category with both tests. However, for QFT-Plus, significance was lost after adjusting for confounding variables. Thus, VIDAS^®^ TB-IGRA may discriminate among persons at different levels of TB-exposure risk.

TB continues to pose a significant threat to public health, and reliable diagnostic methods are needed to support global TB control and eradication efforts [40]. The VIDAS^®^ TB-IGRA represents the first test of its kind that is fully automated from sample input to results interpretation, thereby facilitating ease of use, saving time and improving laboratory workflows, while decreasing the possibility for human error and variability. Our data demonstrates that VIDAS^®^ TB-IGRA has a higher sensitivity for TB infection detection compared to QFT-Plus— the most used IGRA worldwide—suggesting that this new assay can be a tool to improve current TB diagnostic capabilities.

A limitation of this study is that study participants were recruited from only two countries with high TB-incidence rates, South Africa and Mexico. However, the results obtained in this study are similar to those from a previous study [41] with, notably, 101 patients with TB disease from Burkina Faso (a high TB-incidence country) as well as France and the US, showing that VIDAS^®^ TB-IGRA had consistently higher sensitivity than QFT-Plus, with fewer indeterminate results. Nevertheless, our study incorporated an international multi-centre design and inclusion of a large cohort of individuals at varying levels of TB-exposure risk, thereby allowing test evaluation over a wide exposure gradient. Future studies evaluating VIDAS^®^ TB-IGRA accuracy in additional high-burden countries, ease of implementation, and better turnaround time in different operational settings are required.

In conclusion, our findings show that compared to the QFT-Plus, the VIDAS^®^ TB-IGRA offers increased sensitivity and reliability (*i.e*., fewer indeterminate results) for TB infection detection, without sacrificing specificity, thus representing a new and valuable tool in global efforts to end TB.

## Data Availability

All data produced in the present study are available upon reasonable request to the authors

## ACKNOWLEDGEMENTS

This work was funded by bioMérieux. We are grateful to all study participants. We thank all bioMérieux members who took part in this work. We are grateful to the bioMérieux Data Science team for assisting with the statistical evaluation of the data, and the bioMérieux Clinical Affairs team for coordinating the study. The authors thank Laura Marinelli (BioScience Writers, USA) for providing medical writing support, which was funded by bioMérieux (Marcy L’Etoile, France).

