## Supplementary Information for "VIDAS^®^ TB-IGRA accuracy in tuberculosis patients and persons at varying risk of exposure"

*Study sites*

**Site FR01**
Etablissement Français du Sang (EFS), Lyon, Auvergne Rhône-Alpes, France
Yves Mérieux, Francis Camoin, Marie-Christine Lambert, Carine Marliere
Clinical Affairs laboratory, bioMérieux, Marcy l'étoile, France
Laurence Bridon, Pauline Fabre, Aurélie Langlet, Florence Senot, Isabelle Millon

**Site FR02**
Lariboisière Hospital, Paris, France
Dr. Amanda Lopes, Prof Emmanuelle Cambau, Dr Faiza Mougari, Prof Philippe Manivet, Claire Pernin, Lydia Suarez, Valerie Andrianasolonirina, Véronique Delasse

**Site FR03**
Avicenne Hospital, Paris, France
Dr. Frédéric Méchaï, Fadhila Messani, Miassa Slimani, Ingrid Jacquelin. Laboratory (TB culture): Pr. Etienne Carbonnelle, Laure Poulet

**Site FR04**
CHU de Saint-Etienne 42055 Saint-Etienne cedex 02
Prof. Elisabeth Botelho-Nevers.
Infectious diseases: Dr Amandine Gagneux-Brunon, Dr Anne Fresard, Veronique Ronat, Philippine Bourrassaud, Anne Pouvaret. Pneumology: Prof Jean Michel Vergnon, Dr Marios Froudarakis, Rémi Chapelon. Dermatology: Prof Jean Luc Perrot, Marie Chazelle. Rheumatology: Prof Hubert Marotte, Hervé Locrelle. Occupational Medicine: Prof Luc Fontana, Marie Christine Coquet. Laboratory: Dr Claude Lambert, Dr Anne Berger, Alice Haccourt, Alice Chanavat

**Site FR05**
Anti-TB center of Chambéry, Chambéry, France
Dr. Margaux Isnard, Dr Cécile Descotes-Genon, Ms Marie-Christine Carret, Mr Tarik Habet, Dr Marion Levast, Ms Evelyne Capitan, Corinne Bernati

**Site FR06**
Lapeyronie Hospital, Montpellier, France
Dr. Edouard Tuaillon.
Infectious diseases: Quam Edem Aquereburu. Rheumatology: Prof Jacques Morel, Beatrice Tinland, Sylvie Gouilloux. Occupational Medicine: Dr François-Xavier Lesage, Dr Guillaume Choron, Xavier Loche. Laboratory: Dr Amandine Pisoni, Brigitte Burrut, Françoise Bel.

**Site FR07**
Anti-TB center of Nanterre, Nanterre, France
Dr. Amel Medjahed-Artebasse

**Bacteriology laboratory, Antoine Béclère Hospital (APHP), Clamart, France**
*Testing of samples from sites FR03 and FR07*
Pr Doucet- Populaire Florence, Dr Christelle Guillet-Caruba, Rachida Aït El Far, Sandrine Roze, Daniel Martinho

**Site GB01**
St Thomas' Hospital, London, UK
Dr. Ronan Breen, Arbane Gill, Laletha Agoramoorthy

**Site GB02**
Evelina London Children's Hospital, London, UK
Dr. Julia Kenny, Dr Marc Tebruegge, Bethany Hamilton, Christian Henderson, Daniella Hydes

**CIDR Laboratory (Centre for Clinical Infection & Diagnostics Research)**
*VIDAS testing for sites GB01 and GB02*
Dr. Rahul Batra, Amita Patel, Bindi Patel

**Site IT01**
INMI L. Spallanzani, Rome, Italy
Dr. Delia Goletti, Dr Elisa Petruccioli, Dr Gina Gualano, Valentina Vanini, Gilda Cuzzi

**Site ME01**
Autonomous University of Baja California, Mexicali, Mexico
Dr. Julia Dolores Estrada Guzman, Dr Rosa Herrera-Torres, Dr Zuceth Duran-Gomez, Dr Fernanda Cervantes-Lopez, Dr Rene Machado-Contreras, Salvador Josaet Cervantes-Borrego, Yosef Dueñez-Uriarte

**Site US01**
Rutgers University, Newark, NJ (USA)
Dr. Maria Laura Gennaro, Dr Alfred Lardizabal, Deborah Handler, Dr. Natalie Bruiners, Dr. Rahul Ukey, Blas Peixoto, Alberta Onyuka, Ariana Alcaide

**Collaborating hospitals with US01**
Doreen Dutchak RN, and Christian Engell, MD, Newark Beth Israel Medical Center
Mabel La Forgia, RN, and Joseph DePasquale, MD, Jersey City Medical Center

**Site US02**
University of California San Diego, San Diego, CA (USA)
Dr. David T. Pride, Nathan Kendrick, Steven Hendrickx, Stephanie Solso, Helene Le, Joseph Lencioni, Rueben Barba, Maria Pizzaro, Jenny Shin

**Site US04**
Stanford University, Palo Alto, CA (USA)
Dr. Niaz Banaei, Arena Shafeque, Fiona Senchyna

**Site US05**
Saint Louis University, St Louis, MO (USA)
Dr. Daniel Hoft, Dr. Azra Blazevic, Linda Eggemeyer-Sharpe, Janice Tennant, Sabrina Dipiazza, Kate Lierfer, Joan Siegner, Amanda Nethington, Tammy Blevins, Yinyi Yu, Huan Ning

**Site US06**
National Jewish Health, Denver, CO (USA)
Dr. Charles Daley, Dr Robert Belknap, Dr Michael Wilson, Michael Higgins, Kaylynn Aiona

**Site US07**
University of Illinois, Chicago, IL (USA)
Dr Nahed Ismail, Dr Susan Bleasdale, Anh Nguyen, Iuliana Bentea, Brenna Lindsey

**Site ZA01**
University of Cape Town, Cape Town, South Africa
Prof. Keertan Dheda, Dr Aliasgar Esmail, Dr Suzette Oelofse, Dr Lynelle Mottay, Marietjie Pretorius, Richard Meldau, Carley Mandviwala, Maria Gorgel, Eleanor Pretorius, Solomri Foloti, Lusanda Yenani, Vonnita Loren, Wasiela Galant

**Site ZA02**
TASK Applied Science, Cape Town, South Africa
Dr Naadira Vanker, Dr Caryn Upton, Michelle Eriksson, Martha Jacobs, Anastacia Arendse, Wendy Wakens, Dorothy Zakariya, Alexandra Waltman, Chantell Meyer, Margaret Muller, Michele van Rooyen, Phumza Setlaba, Patricia Maunder, Mlungisi Mnunu, Doreen Petersen, Charné Rossouw, Magdalene Kennedy, Silvia Nunes, Thirumani Govender, Lize Greyling, Jacqueline Russouw, Katriena Ross, Tracy Jack, Kurt McQuire, Diane Lavies, Beulah Classen, Carmen Kleinhans, Julia Sims, Bryan Esterhuizen, Sven Friedrich, Cebisa Mdladla, Bongiwe Gwadela, Fay Swanson, Pascal Musoni, Dustin Bosch, Lucille October, Atica Moosa, Karabo Mahlangu, Olwethu Ngxishe, Chaldene Johannes, Chantal Botha, Alta Mwela, Leylani Croy

*Supplementary Methods*

Sensitivity of QFT-Plus and VIDAS® TB-IGRA were independently calculated, as follows:

$$\text{Sensitivity} = 100 * \frac{TP}{TP + FN}$$

where TP = true positives (*i.e.*, culture-confirmed TB disease and positive test result) and FN = false negatives (*i.e.*, culture-confirmed TB disease and negative test result).

To evaluate the VIDAS® TB-IGRA accuracy for the mixed exposure risk population, positive percent agreement (PPA) and negative percent agreement (NPA) relative to the QFT-Plus were calculated as follows, along with the 95% confidence interval (95%CI).

$$\text{PPA} = 100 * \frac{\text{a}}{\text{a} + \text{c}} \quad \text{and} \quad \text{NPA} = 100 * \frac{\text{d}}{\text{d} + \text{b}}$$

where a, b, c, and d are defined as follows:

| | | **QFT-Plus result** | | |
|---|---|---|---|---|
| | | **Positive** | **Negative** | **Total** |
| **VIDAS® TB-IGRA result** | **Positive** | a | b | a+b |
| | **Negative** | c | d | c+d |
| | **Total** | a+c | b+d | a+b+c+d |

*Supplementary Tables and Figures*

**Supplementary Table S1.** Study sites for the TB disease and mixed exposure risk populations.

| Region | Country | Site Code | Site name / City | Principal Investigator | Site responsibilities | TB disease Patients (*n* = 200) | Mixed exposure risk Participants (*n* = 1460) |
|---|---|---|---|---|---|---|---|
| EU | France | FR01* | Clinical Affairs Laboratory, bioMérieux / Marcy l'Etoile | Laurence Bridon | VIDAS testing | – | 125 |
| EU | France | FR02 | Lariboisière Hospital / Paris | Dr. Amanda Lopes | Recruitment, Testing (QFT-Plus and VIDAS) | 5 | – |
| EU | France | FR03** | Avicenne Hospital / Paris | Dr. Frederic Méchaï | Recruitment | 3 | 2 |
| EU | France | FR04 | North Hospital / St. Etienne | Prof. Elisabeth Botelho-Nevers | Recruitment, Testing (QFT-Plus and VIDAS) | 4 | 91 |
| EU | France | FR05 | Anti-TB center of Chambéry | Dr. Margaux Isnard | Recruitment, Testing (QFT-Plus and VIDAS) | 5 | 45 |
| EU | France | FR06 | Lapeyronie Hospital / Montpellier | Dr. Edouard Tuaillon | Recruitment, Testing (QFT-Plus and VIDAS) | – | 86 |
| EU | France | FR07** | Anti-TB center of Nanterre | Dr. Amel Medjahed | Recruitment | – | 127 |
| EU | UK | GB01*** | St Thomas' Hospital / London | Dr. Ronan Breen | Recruitment | 1 | 11 |
| EU | UK | GB02*** | Evelina London Children's Hospital | Dr. Marc Tebruegge replaced by Dr Julia Kenny | Recruitment | – | 3 |
| EU | Italy | IT01 | INMI L. Spallanzani-IRCCS / Rome | Dr. Delia Goletti | Recruitment, Testing (QFT-Plus and VIDAS) | 5 | 28 |
| US | USA | US01 | Rutgers University / Newark | Dr. Maria Laura Gennaro | Recruitment, Testing (QFT-Plus and VIDAS) | 17 | 77 |
| US | USA | US02 | University of California / San Diego | Dr. David T. Pride | Recruitment, Testing (QFT-Plus and VIDAS) | – | 5 |
| US | USA | US04 | Stanford University / Palo Alto | Dr. Niaz Banaei | Recruitment, Testing (QFT-Plus and VIDAS) | – | 144 |
| US | USA | US05 | Saint Louis University | Dr. Daniel Hoft | Recruitment, Testing (QFT-Plus and VIDAS) | – | 207 |
| US | USA | US06 | National Jewish Health / Denver | Dr. Charles Daley | Recruitment, Testing (QFT-Plus and VIDAS) | – | 100 |

| | | | | | | | |
|---|---|---|---|---|---|---|---|
| US | USA | US07 | University of Illinois / Chicago | Dr Nahed Ismail | Recruitment, Testing (QFT-Plus and VIDAS) | – | 179 |
| ROW | Mexico | ME01 | Autonomous University of Baja California / Mexicali | Dr. Julia Dolores Estrada Guzman | Recruitment, Testing (QFT-Plus and VIDAS) | 3 | 77 |
| ROW | South Africa | ZA01 | University of Cape Town | Prof. Keertan Dheda | Recruitment, Testing (QFT-Plus and VIDAS) | 105 | 74 |
| ROW | South Africa | ZA02 | TASK Applied Science / Cape Town | Dr. Naadira Vanker | Recruitment, Testing (QFT-Plus and VIDAS) | 52 | 79 |

** For the additional responsibilities:*

- *Recruitment was performed by the national French organization for blood donation (EFS) Auvergne Rhône-Alpes*
- *QFT®-Plus testing was performed by St Etienne Hospital (site FR04 laboratory)*

*** Testing (QFT-Plus and VIDAS) was performed by Antoine Béclère Hospital (APHP), Bacteriology laboratory, Clamart, France*

**** For the additional responsibilities:*

- *QFT-Plus testing was performed by TDL Trials - The Doctors Laboratory*
- *VIDAS testing was performed by CIDR (Centre for Clinical Infection & Diagnostics Research) Guy's and St Thomas' NHS Foundation Trust, London*

**Abbreviations:** EU, European Union; ROW, Rest of the World; US, United States.

**Supplementary Table S2**. Criteria for stratification of the mixed exposure risk population based on TB-exposure risk level.

<table>
<tr><th colspan="2">TB-exposure risk</th><th>Criteria</th><th colspan="2">Number of individuals (%)</th></tr>
<tr><td rowspan="3">Aggregate Low</td><td>Extremely Low</td><td>Blood donors from low burden countries, without known TB-exposure risk factors</td><td>125 (8.6%)</td><td rowspan="3">333 (22.8%)</td></tr>
<tr><td>Very Low</td><td>Persons without known TB-exposure risk who were screened for TB infection prior to or during immunosuppressive therapy</td><td>90 (6.2%)</td></tr>
<tr><td>Low</td><td>Medical students in low TB-burden countries without known TB-exposure risk factors</td><td>118 (8.1%)</td></tr>
<tr><td rowspan="3">Aggregate Medium</td><td>Medium Low</td><td>Healthcare workers or volunteers without known TB-exposure risk factors</td><td>598 (41.0%)</td><td rowspan="3">679 (46.5%)</td></tr>
<tr><td>Medium</td><td>Persons from TB-related settings in low TB-burden countries</td><td>13 (0.9%)</td></tr>
<tr><td>Medium High</td><td>Immigrants or residents of countries with high TB incidence<br>Persons who spent >1 month in an area with high TB incidence</td><td>68 (4.7%)</td></tr>
<tr><td rowspan="3">Aggregate High</td><td>High</td><td>TB-case contacts (non-household)</td><td>296 (20.3%)</td><td rowspan="3">447 (30.6%)</td></tr>
<tr><td>Very High</td><td>TB-case contacts (same house)</td><td>81 (5.6%)</td></tr>
<tr><td>Extremely High</td><td>TB-case contacts (same bedroom)</td><td>70 (4.8%)</td></tr>
</table>

**Supplementary Table S3**. Institutional Review board (IRB) or Independent Ethics Committee (IEC) approval codes for each study operating site. All sites complied with the Declaration of Helsinki, the International Council for Harmonisation of Technical Requirements for Pharmaceuticals for Human Use (ICH) Good Clinical Practice (GCP) guidelines as applicable to *in vitro* diagnostic (IVD) studies, and the laws and regulations of their respective countries. The Principal Investigator of each site was responsible for ensuring compliance with these regulations and adherence to approved protocols, and written informed consent was obtained from all study participants.

| Site Code | Country | IRB/IEC Code |
|---|---|---|
| FR01 | France | N/A (French Blood Bank) |
| FR02<br>FR03<br>FR04<br>FR05<br>FR06<br>FR07 | France | 2019-A00998-49 |
| GB01<br>GB02 | UK | CTPR01<br>IRAS: 265866; REC: 19/WA/0284<br>CTPR02<br>IRAS: 266217; REC: 19/WA/0285<br>REC: Wales REC 7 |
| IT01 | Italy | INMI "L. Spallanzani" Ethics Committee approval n°35/2019 |
| ME01 | Mexico | 02-01-HGMXL/FMED-UABC-2019-08-29-254 |
| US01 | USA | CTPR01: Pro2019001840; CTPR02: Pro2019001936 |
| US02<br>US05<br>US06 | USA | WIRB Tracking Number<br>CTPR01: 20191965; CTPR02: 20192037; CTPR03: 20192039 |
| US04 | USA | eProtocol#: 53485 |
| US07 | USA | 2019-1145 |
| ZA02 | South Africa | Pharma ethics<br>CTPR01: 190822774; CTPR02: 190822777<br>SAHPRA<br>CTPR01: MD20190803; CTPR02: MD20190804<br>University of Cape Town Human Research Ethics Committee<br>CTPR01 and 2: 840/2019 |

**Abbreviations:** IRAS, Integrated Research Application System; SAHPRA, South African Health Products Regulatory Authority; REC, Research Ethics Committee; WIRB, Western Institutional Review Board.

**Supplementary Table S4**. Cut-off values and result interpretation for the VIDAS® TB-IGRA assay.

| NIL (IU/ml) | AG-NIL (IU/ml) | MIT-NIL (IU/ml) | Interpretation | Clinical Significance |
|---|---|---|---|---|
| < 6.40 | ≥ 0.35 and ≥25% NIL | Any | Positive | *M. tb* infection likely |
| | < 0.35 or ≥ 0.35 and < 25% NIL | ≥ 1.1 | Negative | *M. tb* infection not likely |
| | | < 1.1 | INDETERMINATE MIT Low | Likelihood of *M. tb* infection cannot be determined |
| ≥ 6.40 | Any | | INDETERMINATE NIL High | Likelihood of *M. tb* infection cannot be determined |

**Abbreviations:** AG, antigen; MIT, mitogen (positive control); NIL, negative control.

**Supplementary Table S5.** Negative percent agreement (NPA) and positive percent agreement (PPA) for the VIDAS® TB-IGRA relative to the QFT-Plus test for individuals in the mixed exposure risk population at nine levels of risk for TB, ranging from extremely low to extremely high. NPA and PPA were also calculated for the three main levels of TB exposure: aggregate low, medium, and high.

| TB-Exposure Risk | QFT-Plus Negative results (*n*=1217) | | | | QFT-Plus Positive results (*n*=242) | | | |
|---|---|---|---|---|---|---|---|---|
| | *n* | VIDAS® TB-IGRA | | NPA [95%CI] | *n* | VIDAS® TB-IGRA | | PPA [95%CI] |
| | | Neg. | Pos. | | | Pos. | Neg. | |
| Extremely Low | 119 | 119 | 0 | 100% [96.9, 100.0]% | 6 | 3 | 3 | 50.0% [18.8, 81.2]% |
| Very Low | 84 | 78 | 6 | 92.9% [85.3, 96.7]% | 6 | 4 | 2 | 66.7% [30.0, 90.3]% |
| Low | 117 | 113 | 4 | 96.6% [91.5, 99.1]% | 1 | 1 | 0 | 100.0% [2.5, 100.0]% |
| **Aggregate Low** | 320 | 310 | 10 | 96.9% [94.3, 98.5]% | 13 | 8 | 5 | 61.5% [35.5, 82.3]% |
| Medium Low | 593 | 550 | 43 | 92.7% [90.4, 94.6]% | 5 | 2 | 3 | 40.0% [11.8, 76.9]% |
| Medium | 13 | 12 | 1 | 92.3% [66.7, 98.6]% | 0 | N/A | N/A | N/A |
| Medium High | 31 | 25 | 6 | 80.6% [63.7, 90.8]% | 37 | 34 | 3 | 91.9% [78.7, 97.2]% |
| **Aggregate Medium** | 637 | 587 | 50 | 92.2% [89.8, 94.0]% | 42 | 36 | 6 | 85.7% [72.2, 93.3]% |
| High | 184 | 144 | 40 | 78.3% [71.8, 83.6]% | 112 | 104 | 8 | 92.9% [86.5, 96.3]% |
| Very High | 45 | 36 | 9 | 80.0% [66.2, 89.1]% | 36 | 36 | 0 | 100.0% [90.3, 100.0]% |
| Extremely High | 31 | 20 | 11 | 64.5% [46.9, 78.9]% | 39 | 39 | 0 | 100.0% [91.0, 100.0]% |
| **Aggregate High** | 260 | 200 | 60 | 76.9% [71.4, 81.6]% | 187 | 179 | 8 | 95.7% [91.7, 98.1]% |

**Abbreviations**: CI, confidence interval; QFT-Plus, QuantiFERON Plus.

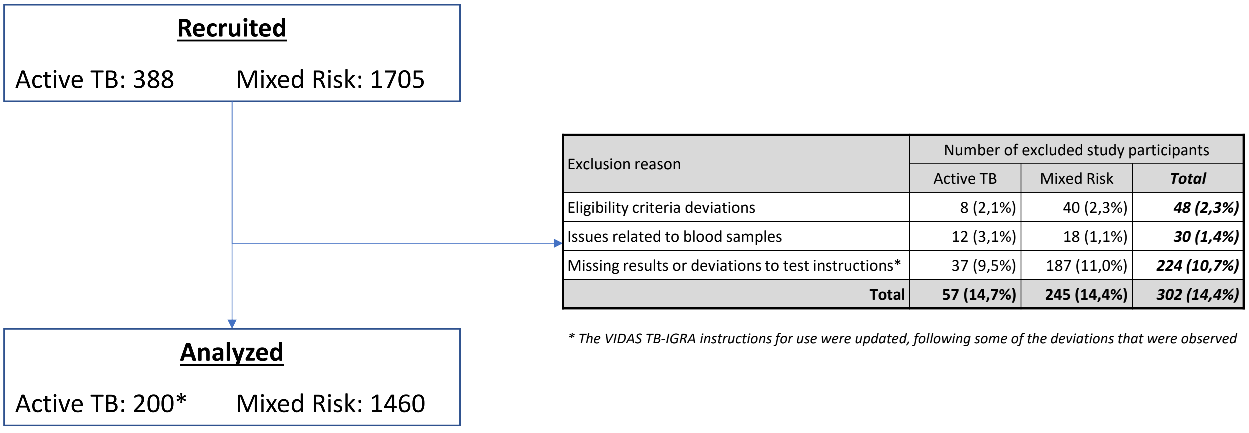


| Exclusion reason | Number of excluded study participants | | |
|---|---|---|---|
| | Active TB | Mixed Risk | ***Total*** |
| Eligibility criteria deviations | 8 (2,1%) | 40 (2,3%) | ***48 (2,3%)*** |
| Issues related to blood samples | 12 (3,1%) | 18 (1,1%) | ***30 (1,4%)*** |
| Missing results or deviations to test instructions* | 37 (9,5%) | 187 (11,0%) | ***224 (10,7%)*** |
| **Total** | **57 (14,7%)** | **245 (14,4%)** | ***302 (14,4%)*** |

** The VIDAS TB-IGRA instructions for use were updated, following some of the deviations that were observed*

** patients not included in sensitivity computations:*

- *76 patients with negative TB-culture*
- *55 patients with positive TB-culture due to sequential design*

**Supplementary Figure S1.** Flow of participants through the study.

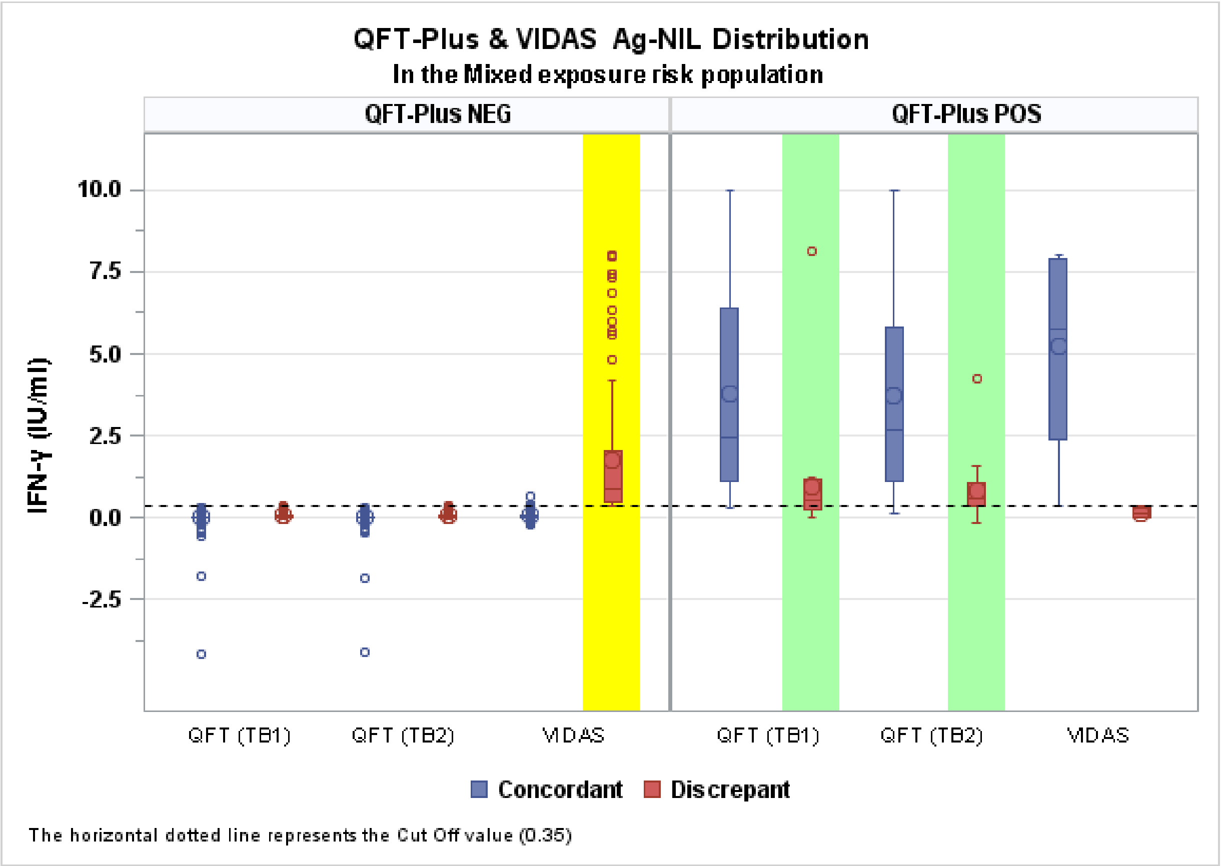


**Supplementary Figure S2.** Distribution of interferon (IFN)-γ concentrations measured by the QFT-Plus and VIDAS® TB-IGRA in the mixed exposure risk population. The dashed line, at 0.35 IU/mL, represents the cut-off that is used for result interpretation. For the VIDAS® TB-

IGRA, the cut-off applies to antigen (AG)-negative control (NIL) value. For the QFT-Plus, the cut-off applies to TB1-NIL and TB2-NIL values. See supplementary table S4 for more details about the VIDAS® TB-IGRA interpretation algorithm. Values from concordant results were spread across the measuring interval for both tests, as follows: i) from 0 to 0.35 IU/mL for concordant negative results (blue boxes on left side of the graph) and ii) from 0.35 IU/mL to the end of the respective measuring interval of each assay, for concordant positive results (blue boxes on right side of the graph). For discrepant results with QFT®-Plus positive (red boxes in green bands), TB1-NIL and TB2-NIL values were grouped more tightly near the cut-off relative to the positive concordant results. In contrast, when the VIDAS® TB-IGRA was positive (red box in yellow band), AG-NIL values covered a large portion of the measuring interval, with several points in the higher half. This suggests that at least a portion of discrepant results were associated with IFN-γ concentrations far from the cut-off, especially when the VIDAS® TB-IGRA was positive.

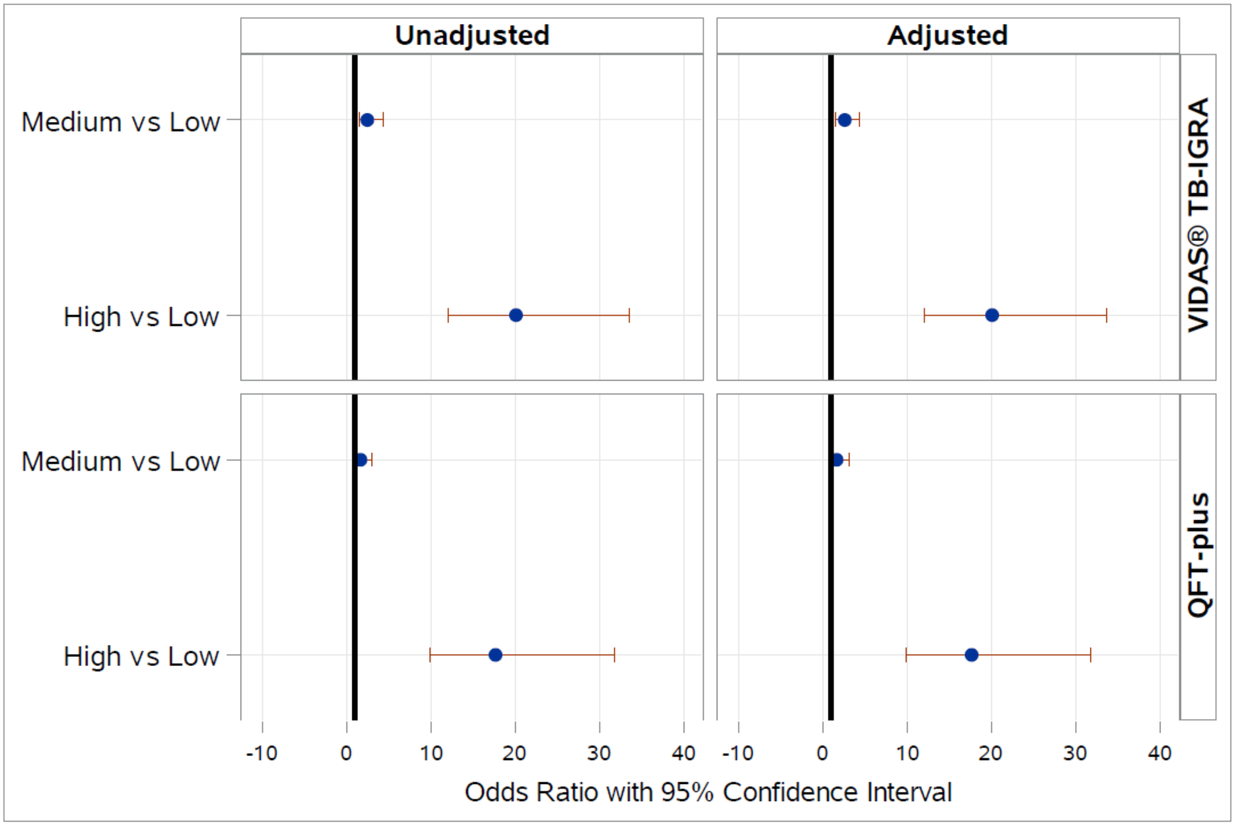


**Supplementary Figure S3.** Univariate and multivariate odds ratios determined by logistic regression for aggregate TB exposure risk groups.

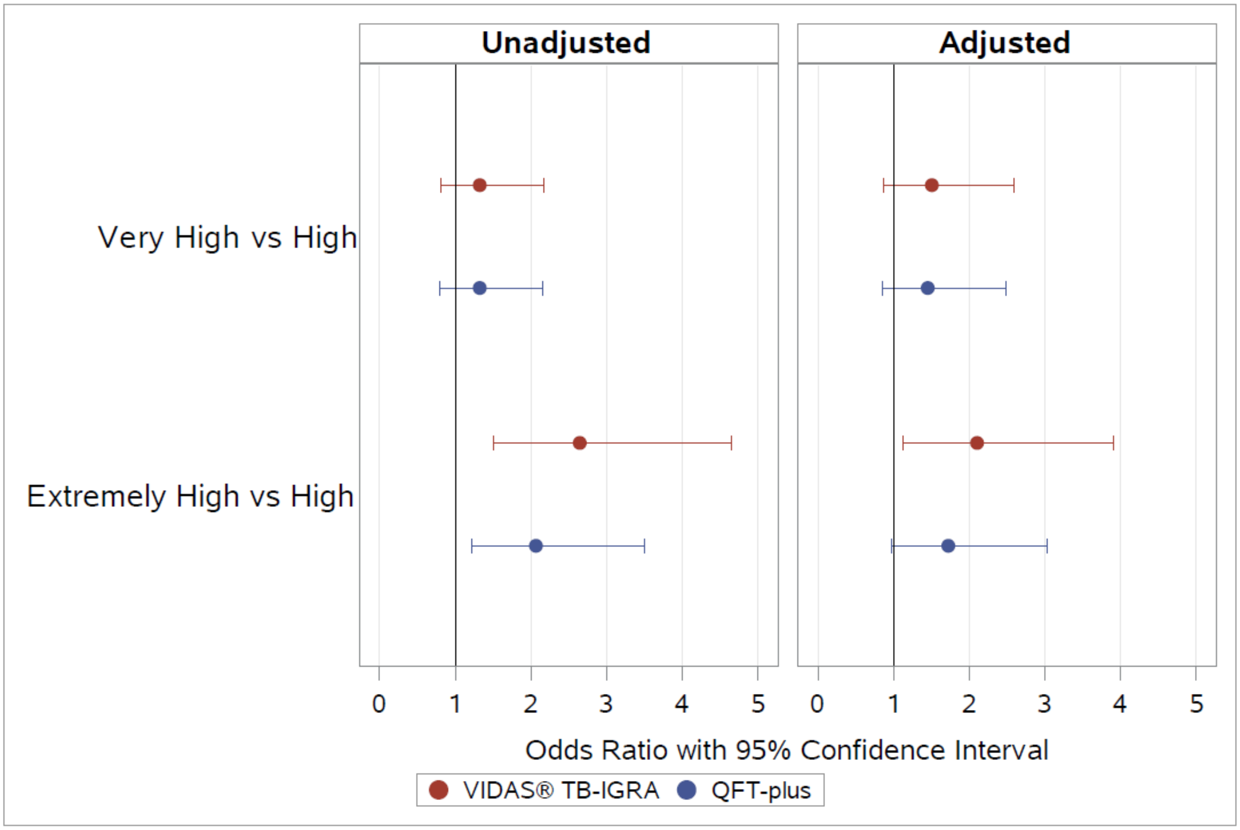


**Supplementary Figure S4.** Univariate and multivariate odds ratios determined by logistic regression for the three subgroups in the aggregate-high TB exposure risk group.
